# Fairness in Cardiac Magnetic Resonance Imaging: Assessing Sex and Racial Bias in Deep Learning-based Segmentation

**DOI:** 10.1101/2021.07.19.21260749

**Authors:** Esther Puyol-Antón, Bram Ruijsink, Jorge Mariscal Harana, Stefan K. Piechnik, Stefan Neubauer, Steffen E. Petersen, Reza Razavi, Phil Chowienczyk, Andrew P King

## Abstract

**Background:** Artificial intelligence (AI) techniques have been proposed for automation of cine CMR segmentation for functional quantification. However, in other applications AI models have been shown to have potential for sex and/or racial bias.

**Objectives:** To perform the first analysis of sex/racial bias in AI-based cine CMR segmentation using a large-scale database.

**Methods:** A state-of-the-art deep learning (DL) model was used for automatic segmentation of both ventricles and the myocardium from cine short-axis CMR. The dataset consisted of end-diastole and end-systole short-axis cine CMR images of 5,903 subjects from the UK Biobank database (61.5±7.1 years, 52% male, 81% white). To assess sex and racial bias, we compared Dice scores and errors in measurements of biventricular volumes and function between patients grouped by race and sex. To investigate whether segmentation bias could be explained by potential confounders, a multivariate linear regression and ANCOVA were performed.

**Results:** We found statistically significant differences in Dice scores (white ∼94% vs minority ethnic groups 86-89%) as well as in absolute/relative errors in volumetric and functional measures, showing that the AI model was biased against minority racial groups, even after correction for possible confounders.

**Conclusions:** We have shown that racial bias can exist in DL-based cine CMR segmentation models. We believe that this bias is due to the imbalanced nature of the training data (combined with physiological differences). This is supported by the results which show racial bias but not sex bias when trained using the UK Biobank database, which is sex-balanced but not race-balanced.

**Condensed Abstract:** AI algorithms have the potential to reflect or exacerbate racial/sex disparities in healthcare. We aimed to determine the impact of sex and race on the performance of an AI segmentation model for automatic CMR quantification in a cohort of 5,903 subjects from the UK Biobank database, which is sex-balanced but not race-balanced. We tested the model’s bias in performance using Dice scores and absolute/relative errors in measurements of biventricular volumes and function. Our study demonstrates that the model had a racial bias but no sex bias, and that subject characteristics and co-morbidities could not explain this bias.

## 1. Introduction

Artificial intelligence (AI) is a rapidly evolving field in medicine, especially cardiology. AI has the potential to aid cardiologists in making better decisions, improving workflows, productivity, cost-effectiveness, and ultimately patient outcomes (1). Deep learning (DL) is a recent advance in AI which allows computers to learn a task using data instead of being explicitly programmed. Several studies in cardiology and other applications have shown that DL methods can match or even exceed human experts in tasks such as identifying and classifying disease (2–4).

In cardiology, cardiovascular imaging has a pivotal role in diagnostic decision making. Cardiac magnetic resonance (CMR) is the established non-invasive gold-standard modality for quantification of cardiac volumes and ejection fraction (EF). For decades, clinicians have been relying on manual or semi-automatic segmentation approaches to trace the cardiac chamber contours. However, manual expert segmentation of CMR images is tedious, time-consuming and prone to subjective errors. Recently, DL models have shown remarkable success in automating many medical image segmentation tasks. In cardiology, human-level performance in segmenting the main structures of the heart has been reported (5, 6), and researchers have proposed to use these models for tasks such as automating cardiac functional quantification (7). These methods are now starting to move towards broader clinical translation.

In the vast majority of cardiovascular diseases (CVDs), there are known associations between sex/race and epidemiology, pathophysiology, clinical manifestations, effects of therapy, and outcomes (8–10). Furthermore, in clinically asymptomatic individuals the Multi-Ethnic Study of Atherosclerosis (MESA) study showed that men had greater right ventricular (RV) mass and larger RV volumes than women, but had lower RV ejection fraction; African-Americans had lower RV mass than whites, whereas Hispanics had higher RV mass (11); and the LV was more trabeculated in African-American and Hispanic participants than white participants, and smoothest in Chinese-American participants (12), but the greater extent of LV trabeculation was not associated with an absolute decline in LVEF during the approximately 10 years of the MESA study. Similarly, the Coronary Artery Risk Development in Young Adults (CARDIA) study (13) showed differences between races (African American and white) and sexes in LV systolic and diastolic function, which persist after adjustment for established cardiovascular risk factors.

Although these physiological differences are associations and not proven causative links with race/gender, their presence raises a potential concern about the performance of AI models in cardiovascular imaging. Although AI has great potential in this area, no previous work has investigated the fairness of such models. In AI, the concept of ‘fairness’ refers to assessing AI algorithms for potential bias based on demographic characteristics such as race and sex. In general, AI models are trained agnostic to demographic characteristic, and they assume that if the model is unaware of these characteristics while making decisions, the decisions will be fair. However, we have recently shown, for the first time, that using this assumption there exists racial bias in DL-based cine CMR segmentation models when trained using racially imbalanced data (14). That preliminary work focused on the technical development of different bias mitigation strategies, in order to reduce the bias effect between different racial groups. The object of this study is to investigate in more detail the origin and the effect of this bias on cardiac structure and function using a standard AI segmentation model, and also to assess whether it can be explained by other possible confounders such as subject characteristics or cardiovascular risk factors.

## 2. Methods

### 2.1. Participants

The UK Biobank is a prospective cohort study with more than 500,000 participants aged 40 to 69 years of age conducted in the United Kingdom (15). This study complies with the Declaration of Helsinki; the work was covered by the ethical approval for UK Biobank studies from the NHS National Research Ethics Service on 17th June 2011 (Ref 11/NW/0382) and extended on 18 June 2021 (Ref 21/NW/0157) with written informed consent obtained from all participants. The present study was performed using a sub-cohort of the UK Biobank imaging database, for whom CMR imaging and ground truth manual segmentations were available. In this study, in order to minimise the effects of physiological differences due to cardiovascular and other related diseases, we only focus on the healthy population of the UK Biobank database and analyse possible confounders that can explain racial and sex bias. Therefore, we excluded any subjects with known cardiovascular disease, respiratory disease, haematological disease, renal disease, rheumatic disease, malignancies, symptoms of chest pain, respiratory symptoms or other diseases impacting the cardiovascular system, except for diabetes mellitus, hypercholesterolemia and hypertension (see all exclusion criteria in Supplementary List 1). We used the ICD-9 and ICD-10 codes and self-reported detailed health questionnaires and medication history for the selection process.

In this paper, race was assumed to align with self-reported ethnicity, which was the data collected in the UK Biobank. From the total UK Biobank database (N=501,642), the race distribution is as follows: White 94.3%, Mixed 0.6%, Asian, 1.9%, Black 1.6%, Chinese: 0.9%, Other: 0.4%. The UK Biobank cohort has a similar ethnic distribution to the national population of the same age range in the 2011 UK Census^1^. The imaging cohort used in this study (N=5,660) has a slightly different racial distribution (White 81%, Mixed 3%, Asian, 7%, Black 4%, Chinese: 2%, Other: 3%), but it is still predominantly White race, in line with the full cohort of the UK Biobank database.

Subject characteristics obtained were age, binary sex category, race, body measures (height; weight; body mass index, BMI; and body surface area, BSA), and smoker status (smoker was defined as a subject smoking or smoked daily for over 25 years in the previous 35 years). We also obtained the average heart rate (HR) and brachial systolic and diastolic blood pressure (SBP and DBP) measured during the CMR exam. These subject characteristics were considered as possible confounders in the statistical analysis.

### 2.2. Automated image analysis

A state-of-the-art DL based segmentation model, the ‘nnU-Net’ framework (16), was used for automatic segmentation of the left ventricle blood pool (LVBP), left ventricular myocardium (LVMyo) and right ventricle blood pool (RVBP) from cine short-axis CMR slices at end-diastole (ED) and end-systole (ES). This model was chosen as it has performed well across a range of segmentation challenges and was the top-performing model in the ‘ACDC’ CMR segmentation challenge (6). For training and testing the segmentation model, we used a random split with similar sex and racial distributions of 4,410 and 1,250 subjects respectively. We refer the reader to our previous paper (14) for further details of the model architecture and training.

#### Evaluation of the method

For quantitative assessment of the image segmentation model, we used the dice similarity coefficient (DSC), which quantifies the overlap between an automated segmentation and a ground truth segmentation. DSC has values between 0 and 100%, where 0 denotes no overlap, and 100% denotes perfect agreement. From the manual and automated image segmentations, we calculated the LV end-diastolic volume (LVEDV) and end-systolic volume (LVESV), and RV end-diastolic volume (RVEDV) and end-systolic volume (RVESV) by summing the number of voxels belonging to the corresponding label classes in the segmentation and multiplying this by the volume per voxel. The LV myocardial mass (LVmass) was calculated by multiplying the LV myocardial volume by a density of 1.05 g/mL. Derived from the LV and RV volumes, we also computed LV ejection fraction (LVEF) and RV ejection fraction (RVEF). We evaluated the accuracy of these volumetric and functional measures by computing the absolute and relative differences between automated and manual measurements. We define the absolute and relative error as *ε*_*absolute*_ = | *ν*_*manual*_ − *ν*_*auto*_|) and *ν*_*relative*_(%) = 100*| *ν*_*manual*_ − *ν*_*auto*_|/*ν*_*manual*_, where *ν* corresponds to each clinical measure.

### 2.3. Analysis of the influence of confounders

To investigate whether a true bias between racial and/or sex groups exists for automated DL-based cine CMR segmentation, we conducted a statistical analysis to investigate if the observed bias could be explained by the most common confounders. In this study, we use as possible confounders age, sex, body measures (i.e. height, weight and BMI), HR, SBP, DBP, CMR-derived parameters (HR, LVEDV, LVESV, RVEDV, RVESV, LVmass), cardiovascular risk factors (i.e. hypertension, hypercholesteremia, diabetes and smoking) and centre (i.e. core lab where most of the segmentations were performed versus additional lab).

### 2.4. Statistical analysis

Data analysis was performed using SPSS Statistics (version 27, IBM, USA). Continuous variables are reported as mean ± standard deviation (SD) and tested for normal distributions with the Shapiro–Wilk test. Log transformations were applied to the (1-DSC) values to obtain an approximately normal distribution. After transformation, all continuous variables were normally distributed. Categorical data are presented as absolute counts and percentages. Comparison of variables between groups (i.e. races and sex) was carried out using an independent Student’s *t-*test. Pair-wise post hoc testing was carried out using Scheffé correction for multiple comparisons.

Independent association between log-transformed DSC values and race was performed using univariate linear regression followed by multivariate adjustment for confounders. All variables in the regression models were standardised by computing the z-score for individual data points. Finally, the differences in DSC values among different racial groups were initially assessed by a 1-way ANOVA (Model 4) followed by an analysis of covariance – ANCOVA (Model 5) to statistically control the effect of covariates. For all statistical analysis, the threshold for statistical significance was p<0.01 and confidence intervals (%) were calculated by non-parametric bootstrapping with 1000 resamples.

## 3. Results

### 3.1. Subject characteristics

The dataset used consisted of ED and ES short-axis cine CMR images of 5,660 healthy subjects (with or without cardiovascular risk factors). Subject characteristics for all participants were obtained from the UK Biobank database and are provided in Table 1.

**Table 1:**
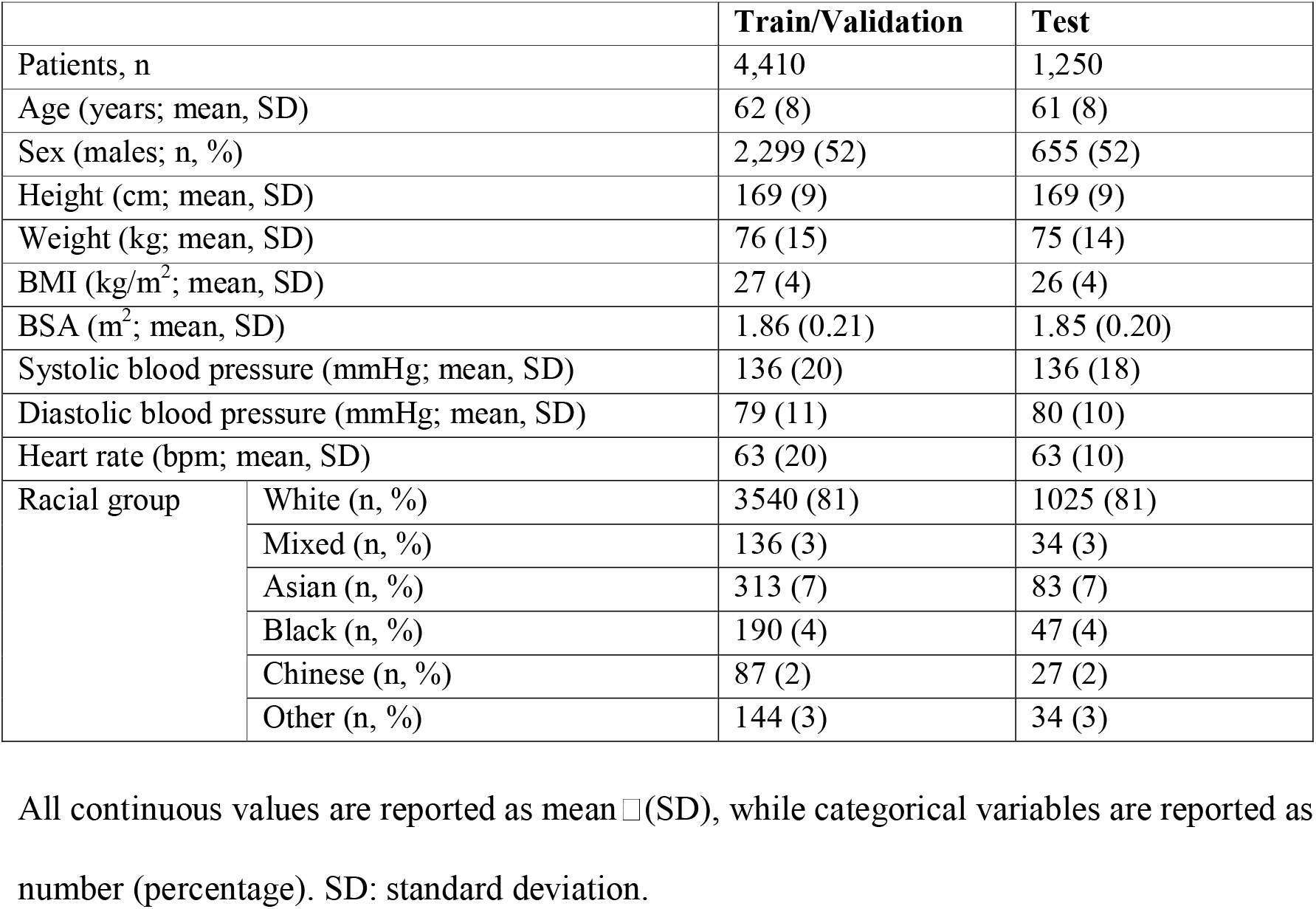
Population characteristics for the train/validation and test sets.

|  |  | **Train/Validation** | **Test** |
| --- | --- | --- | --- |
| Patients, n |  | 4,410 | 1,250 |
| Age (years; mean, SD) |  | 62 (8) | 61 (8) |
| Sex (males; n, %) |  | 2,299 (52) | 655 (52) |
| Height (cm; mean, SD) |  | 169 (9) | 169 (9) |
| Weight (kg; mean, SD) |  | 76 (15) | 75 (14) |
| BMI (kg/m<sup>2</sup>; mean, SD) |  | 27 (4) | 26 (4) |
| BSA (m<sup>2</sup>; mean, SD) |  | 1.86 (0.21) | 1.85 (0.20) |
| Systolic blood pressure (mmHg; mean, SD) |  | 136 (20) | 136 (18) |
| Diastolic blood pressure (mmHg; mean, SD) |  | 79 (11) | 80 (10) |
| Heart rate (bpm; mean, SD) |  | 63 (20) | 63 (10) |
| Racial group | White (n, %) | 3540 (81) | 1025 (81) |
|  | Mixed (n, %) | 136 (3) | 34 (3) |
|  | Asian (n, %) | 313 (7) | 83 (7) |
|  | Black (n, %) | 190 (4) | 47 (4) |
|  | Chinese (n, %) | 87 (2) | 27 (2) |
|  | Other (n, %) | 144 (3) | 34 (3) |

4 All continuous values are reported as mean ± (SD), while categorical variables are reported as
5 number (percentage). SD: standard deviation.

For all subjects, the LV endocardial and epicardial borders and the RV endocardial border were manually traced at ED and ES frames using the cvi42 software (version 5.1.1, Circle Cardiovascular Imaging Inc., Calgary, Alberta, Canada). 4,975 subjects were previously analysed by two core laboratories based in London and Oxford (17), the remaining 685 subjects were analysed by two experienced CMR cardiologists at Guy’s and St Thomas’ Hospital following the same standard operating procedures described in (17). For all CMR examinations that underwent manual image analysis, any case with insufficient quality (i.e. presence of artefacts or slice location problems, operator error or evidence of pathology, such as significant shunt or valve regurgitation) were rejected (18). All experts performing the segmentations were blinded to subject characteristics such as race and sex. From our database, 4,410 subjects were used to train the DL-based CMR segmentation model, and 1,250 subjects were used as a test set for the validation of the model and the statistical analysis. The train and test sets were stratified to contain approximately the same percentage of samples for each racial group and sex.

### 3.2. DL-based image segmentation pipeline

Table 2 reports the DSC values between manual and automated segmentations evaluated on the test set of 1,250 subjects which the segmentation model had never seen before. The table shows the mean DSC for LVBP, LVMyo and RVBP for both the full test set and stratified by sex and race. Overall, the average (AVG) DSC was 93.03±3.83% (94.40±2.61% for the LVBP, 88.78±3.08% for the LVMyo and 90.77±3.96% for the RVBP). Table 2 shows that the CMR segmentation model had a racial bias for all comparisons but no sex bias (independent Student’s *t-*test between each racial group and rest of the population; p < 0.001 for LVBP, LVMyo, RVBP and AVG for all races).

**Table 2:**
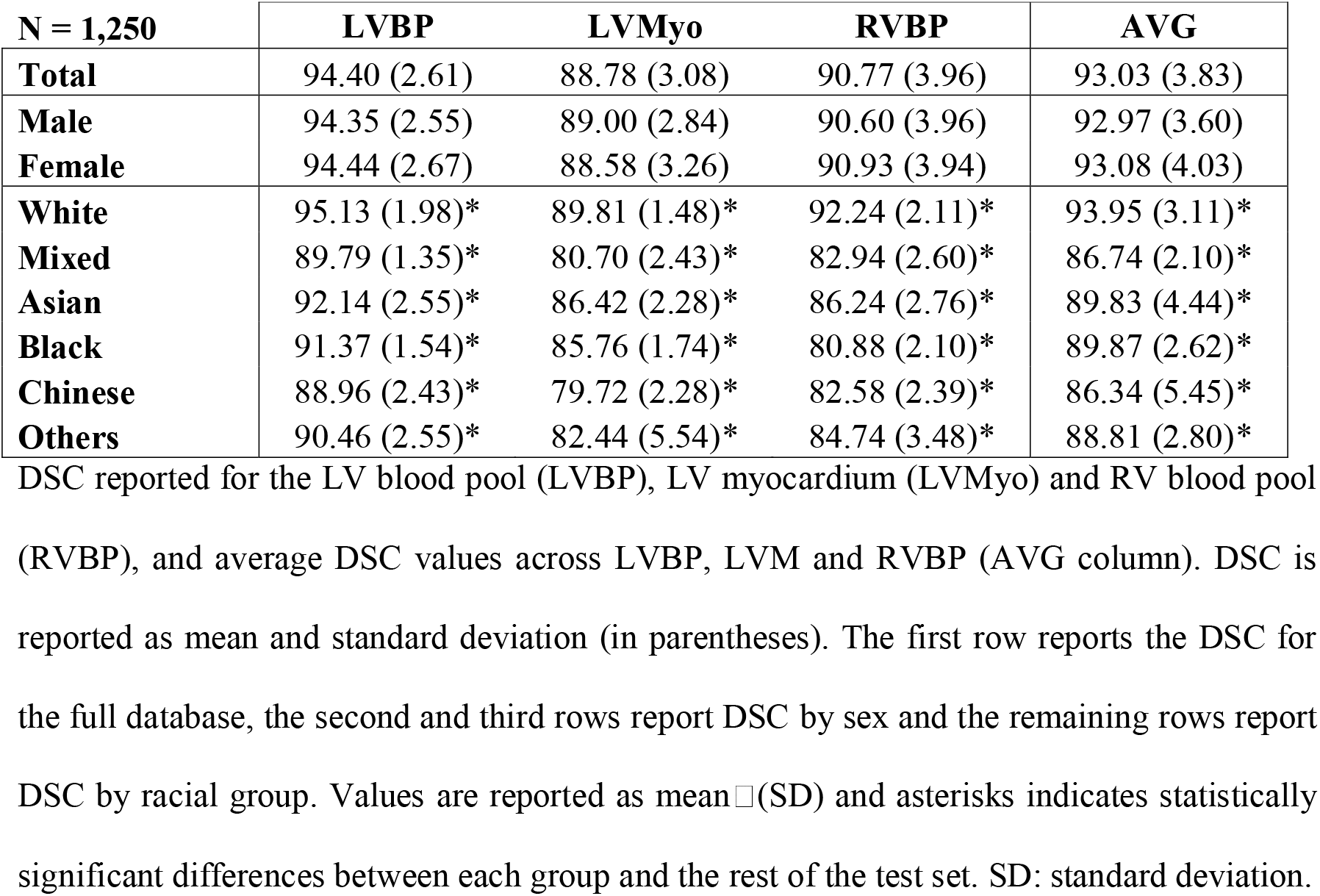
Dice similarity coefficient (DSC) values for the overall test set and by sex and race.

| **N = 1,250** | **LVBP** | **LVMyo** | **RVBP** | **AVG** |
| --- | --- | --- | --- | --- |
| **Total** | 94.40 (2.61) | 88.78 (3.08) | 90.77 (3.96) | 93.03 (3.83) |
| **Male** | 94.35 (2.55) | 89.00 (2.84) | 90.60 (3.96) | 92.97 (3.60) |
| **Female** | 94.44 (2.67) | 88.58 (3.26) | 90.93 (3.94) | 93.08 (4.03) |
| **White** | 95.13 (1.98)* | 89.81 (1.48)* | 92.24 (2.11)* | 93.95 (3.11)* |
| **Mixed** | 89.79 (1.35)* | 80.70 (2.43)* | 82.94 (2.60)* | 86.74 (2.10)* |
| **Asian** | 92.14 (2.55)* | 86.42 (2.28)* | 86.24 (2.76)* | 89.83 (4.44)* |
| **Black** | 91.37 (1.54)* | 85.76 (1.74)* | 80.88 (2.10)* | 89.87 (2.62)* |
| **Chinese** | 88.96 (2.43)* | 79.72 (2.28)* | 82.58 (2.39)* | 86.34 (5.45)* |
| **Others** | 90.46 (2.55)* | 82.44 (5.54)* | 84.74 (3.48)* | 88.81 (2.80)* |

DSC reported for the LV blood pool (LVBP), LV myocardium (LVMyo) and RV blood pool (RVBP), and average DSC values across LVBP, LVM and RVBP (AVG column). DSC is reported as mean and standard deviation (in parentheses). The first row reports the DSC for the full database, the second and third rows report DSC by sex and the remaining rows report DSC by racial group. Values are reported as mean ± (SD) and asterisks indicates statistically significant differences between each group and the rest of the test set. SD: standard deviation.

Next, we evaluate the accuracy of the volumetric and functional measures (LVEDV, LVESV, LVEF, LVmass, RVEDV, RESV, RVEF). Table 3(a) reports the mean values based on the manual segmentations, and Table 3(b) and 3(c) report the mean absolute differences and relative differences between automated and manual measurements, respectively. For the overall population, results are in line with previous reported values (5, 19) and within the inter-observability range (17).

**Table 3:**
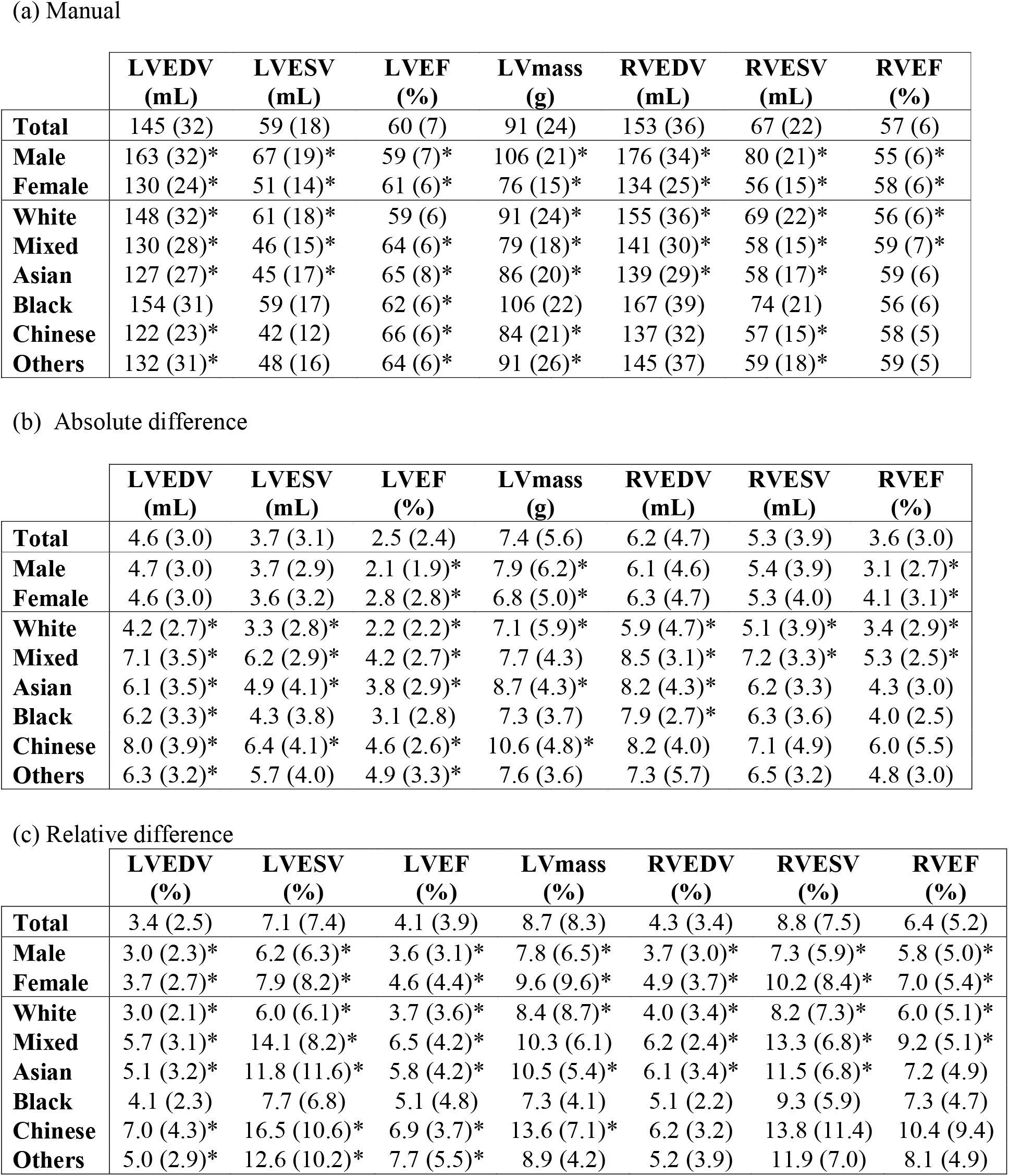
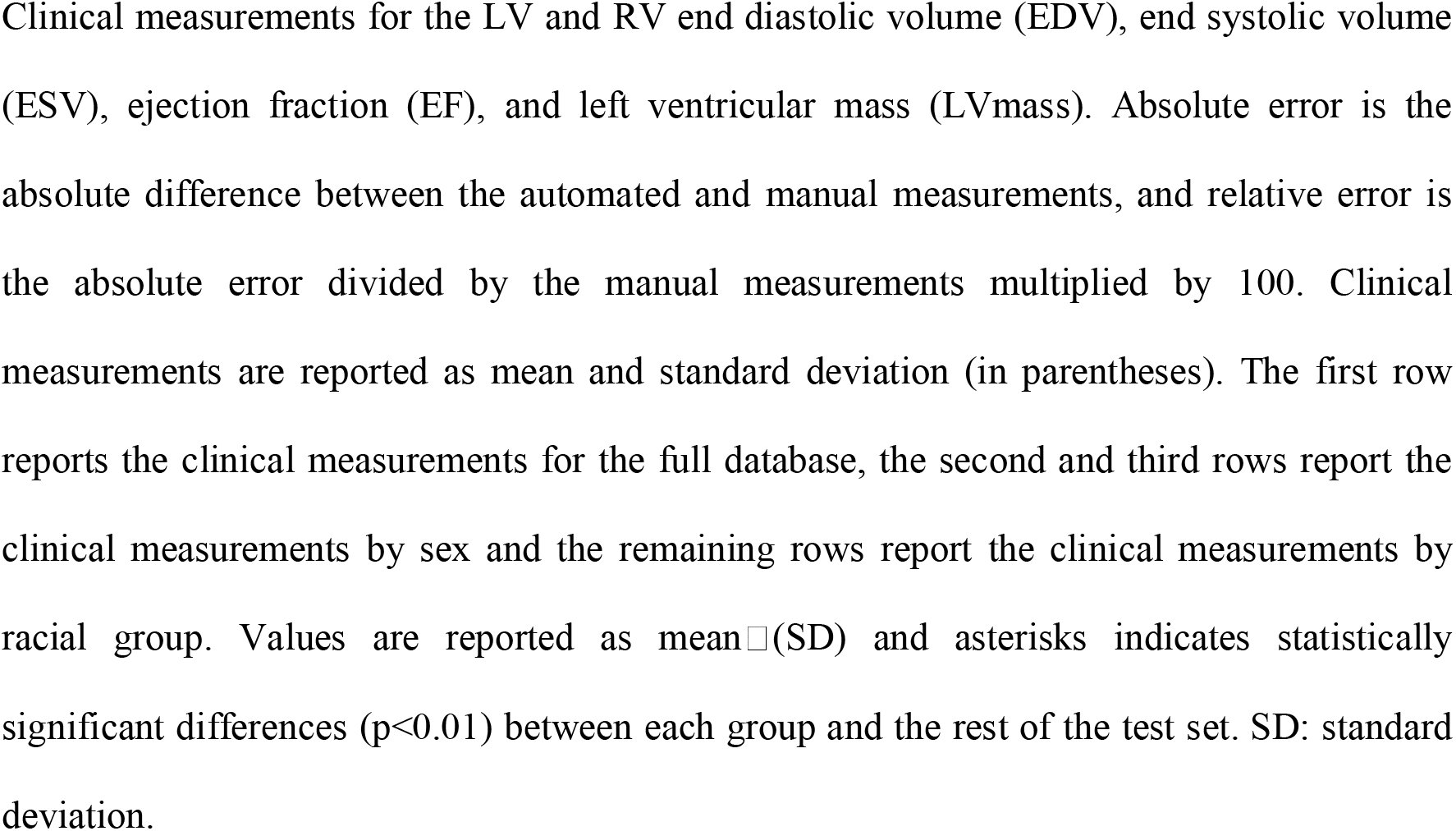
Manual clinical measurements (top table) and absolute (middle table) and relative (bottom table) differences in volumetric and functional measures between automated and manual segmentations, overall and by sex and race.

These results show that for sex there is a statistically significant difference in the absolute error for LVEF, LVmass and RVEF (independent Student’s *t-*test p < 0.001). For different racial groups, they show that the White and Mixed groups have for all clinical parameters a statistically significant difference in absolute and relative error (except Mixed LVmass p=0.66 and p=0.15 for absolute and relative error respectively). They also show that there is a statistically significant difference in the absolute and relative errors for LVEDV, LVESV, LVEF (except for absolute error for Black and Other LVESV p=0.25 and p=0.01 respectively, and Black LVEF p=0.17; and relative error for Black LVEDV p=0.03, LVESV p=0.53 and LVEF p=0.20). Interestingly, there is no statistically significant difference in absolute or relative error for RV clinical parameters for the Chinese and Other racial groups.

### 3.3. Multivariable analysis

To analyse if there is any other factor (i.e. risk factors, patient characteristics) that could explain the bias in DSC between races, we performed a multivariate linear regression between the DSC and race adjusted for patient size, cardiac parameters and cardiovascular risk factors. Table 4 shows the unadjusted (model 1 – 4(a)) and adjusted (model 2 – 4(b)) standardized regression beta coefficients (with 95% confidence interval (CI)) for the association between DSC and racial groups. Supplementary Table 2 shows the full list of standardized regression beta-coefficients from the multivariate analysis for each racial group (model 3), representing the z-score change in variables with the associated factors. Our results show that all associations remained significant after multivariate adjustment and that there is no covariate that can explain the DSC bias between racial groups (see Table 4(b)). For the Mixed and Black race groups, sex shows a weak positive association with DSC (see Supplementary Table 2), however, race remains the main factor.

**Table 4:**
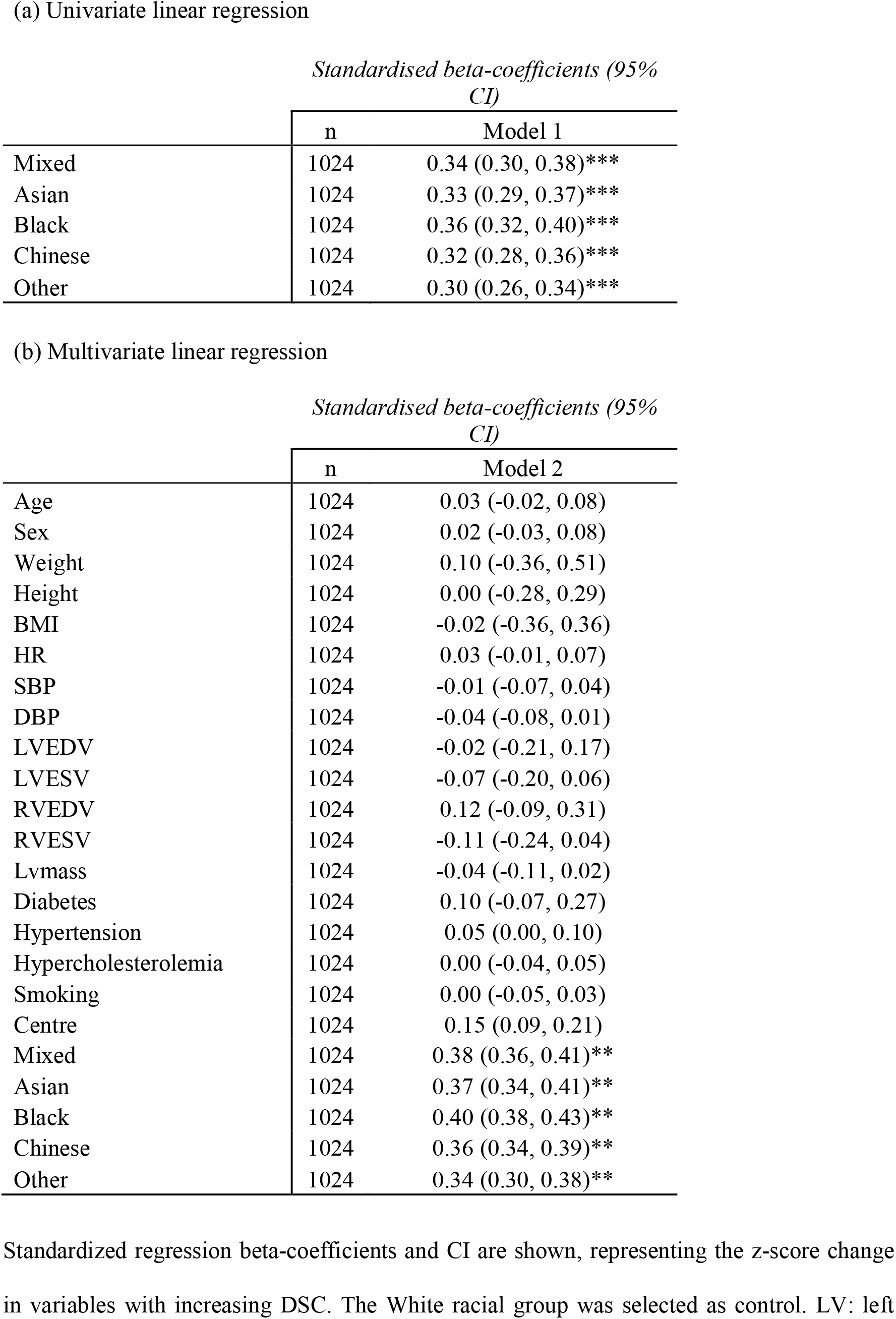
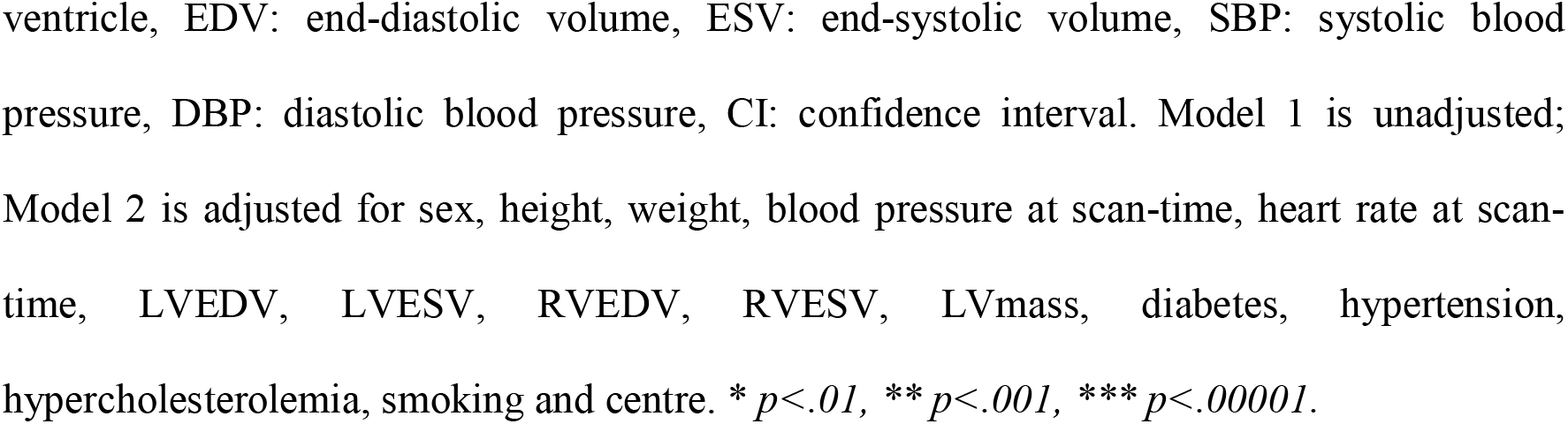
Associations between average DSC and racial group.

### 3.4. Analysis of variance

We also compared change of marginal means of DSC between different racial groups using a 1-way ANOVA (*F* = 219.43, *p* < 0.0001, *η*^2^ = 0.47) and an ANCOVA adjusted for patient size, cardiac parameters and cardiovascular risk factors (*F* = 196.237, < 0.0001, *η*^2^ = 0.44), see Supplementary Table 3. Estimated marginal means are given and displayed in Table 5, before and after adjustment for the mean of covariates. Results show that there is an overall difference between racial groups, and after adjustment for covariates race still remains the main factor.

**Table 5:** The comparison of adjusted mean between racial groups based on one-way ANOVA and ANCOVA.

|  | Mean (95% CI) |  |  |
| --- | --- | --- | --- |
|  | n | Model 4 | Model 5 |
| White | 1024 | 0.93 (0.93, 0.93) | 0.93 (0.93, 0.93) |
| Mixed | 34 | 0.84 (0.86, 0.82) | 0.83 (0.85, 0.80) |
| Asian | 83 | 0.89 (0.90, 0.88) | 0.88 (0.89, 0.88) |
| Black | 47 | 0.86 (0.87, 0.85) | 0.85 (0.86, 0.83) |
| Chinese | 27 | 0.84 (0.86, 0.81) | 0.82 (0.84, 0.78) |
| Other | 34 | 0.86 (0.88, 0.85) | 0.85 (0.87, 0.83) |

Model 4 is unadjusted; Model 5 is adjusted for sex, height, weight, blood pressure at scan-time, heart rate at scan-time, LVEDV, LVESV, RVEDV, RVESV, LVmass, diabetes, hypertension, hypercholesterolemia, smoking and centre. CI: confidence interval.

### 3.5. Effect of bias on HF diagnosis

The previous experiments have demonstrated that racial bias exists in the DL-based CMR segmentation model. This final experiment aims to provide an example of how this racial bias could potentially have an effect on the diagnosis and characterization of heart failure (HF). To this end, we trained another nnU-Net segmentation model using both healthy and cardiomyopathy subjects from the UK Biobank (training and validation: 4,410 healthy subjects/200 cardiomyopathy subjects and test: 1,250 healthy subjects/150 cardiomyopathy subjects). For the cardiomyopathy test cases, we computed the misclassification rate – MCR (%) between the manual LVEF and the automated LVEF based on the standard classification of HF according to LVEF (20, 21), i.e. HF with reduced EF (HFrEF): HF with an LVEF of ≤40%; HF with mildly reduced EF (HFmrEF): HF with an LVEF of 41% to 49%; HF with preserved EF (HFpEF): HF with an LVEF of ≥50%. The results are presented in Table 6. Overall, although the number of subjects in the minority racial groups was relatively small, the misclassification rate using the AI-derived segmentations for White subjects was low, with generally much higher rates for minority races.

**Table 6:** Misclassification rate for HF diagnosis.

|  | n | HFrEF<br>LVEF < 40% |  | HFmrEF<br>LEF 40-49% |  | HFpEF<br>LVEF >= 50% |  |
| --- | --- | --- | --- | --- | --- | --- | --- |
|  |  | n GT | MCR | n GT | MCR | n GT | MCR |
| White | 107 | 5 | 3.74 | 14 | 5.61 | 88 | 7.48 |
| Mixed | 11 | 3 | 45.45 | 0 | - | 8 | 36.36 |
| Black | 8 | 0 | - | 4 | 12.05 | 4 | 25.00 |
| Asian | 14 | 4 | 21.43 | 2 | 7.14 | 8 | 14.29 |
| Chinese | 4 | 0 | - | 2 | 25.00 | 2 | 50.00 |
| Other | 6 | 1 | 33.33 | 5 | 16.67 | 0 | - |
| Minority groups | 43 | 8 | 23.26 | 13 | 9.30 | 22 | 23.26 |

The table summarizes numbers of subjects in each racial group and HF diagnosis (i.e. HFrEF, HFmrEF and HFpEF), as well as the misclassification rate (MCR) for each racial group and diagnosis. The row Minority groups combines data from the Mixed, Black, Asian, Chinese and Other group. The left column (n overall) shows the number of subjects for each racial group used to compute the MCRs. For each HF diagnosis, the first column shows the number of ground truth positive subjects in that group, and the second column shows the MCR. When computing the MCRs, the ground truth negative subjects were all subjects from the other HF diagnoses for that racial group. HFrEF: HF with reduced EF, HFmrEF: HF with moderate EF, HFpEF: HF with preserved EF. Blank cells show regions with missing data.

## 4. Discussion

We have demonstrated for the first time the existence of racial bias in DL-based cine CMR segmentation. Results show that after adjustment for possible confounders such as cardiovascular risk factors the bias persists, suggesting that it is related to the balance of the database used to train the DL model. This conclusion is supported by our earlier work (14), where a model trained with a (much smaller) racially balanced database had much reduced bias (although poorer performance overall due to the smaller training database).

### 4.1. Assessment of the bias in the DL-based CMR segmentation model

For the overall population, the DSC values are in line with previous reported values (5, 19) and with the inter-observer variability range (17). DSC as well as absolute differences and relative differences show a higher bias on the RV, however, this is expected as previous studies have highlighted the difficulty in manual contouring of the RV and the higher variability between observers (17).

The bias we found in segmentation model performance was near-exclusively based on race. Statistically significant differences in some derived volumetric/functional measures (see Table 3) were found by sex but these differences were small compared to the differences observed in both DSC (Table 2) and volumetric/functional measures (Table 3) by race. Therefore, none of the confounders used in this study could explain the differences by race. Similarly to the complete UK Biobank database, the subcohort that we used is approximately sex-balanced but not race-balanced, and the highest errors were found for relatively underrepresented racial groups. This phenomenon has been observed before in applications in computer vision (22) and medical imaging (23), but never before reported in CMR image analysis.

We believe that this bias is due to the imbalanced nature of the training data. Combined with previous studies that have shown race-based associations with differences in cardiac physiology using diverse databases (10, 11), the imbalance causes the performance of the DL model to be biased towards the physiology of the majority group (i.e. white subjects), to the detriment of performance on minority racial groups.

Our last experiment showed that using the AI-based predicted EF values will result in higher misclassification rates for the minority races compared to the White subjects, which is in line with the other experiments showing a higher bias for the minority groups.

### 4.2. Consistent reporting of sex and racial subgroups in AI models

It is envisioned that AI will dramatically change the way doctors practise medicine. In the short term, it will assist physicians with easy tasks, such as automating measurements, making predictions based on big data, and putting clinical findings into an evidence-based context. In the long term, it has the potential to significantly optimize patient care, reduce costs, and improve outcomes.

With AI models now starting to be deployed in the real world it is essential that the benefits of AI are shared equitably according to race, sex and other demographic characteristics. It has long been known that current medical guidelines have the potential for sex/racial bias due to the imbalanced nature of the cohorts upon which they were based (24, 25). One could think that AI can solve such problems, as they are “neutral” or “blind” to characteristics such as sex and race. However, as we have shown in this paper, when AI models are used naively, they can inherit the bias present in clinical databases. It is important to highlight the shortcomings of AI at this stage before AI models become more widely deployed in clinical practice.

For these reasons, we believe that it is necessary that new standards are established to ensure equality between demographic groups in AI model performance, and that there is consistent and rigorous reporting of performances for new AI models that are intended to be integrated into clinical practice. Similar to (26), we would recommend that any new AI-based publication include a report of performance across a range of demographic subgroups, particularly race/sex.

### 4.3. Strategies to reduce racial bias

The obvious way to mitigate bias due to imbalanced datasets (whether in current clinical guidelines or AI models) is to use more balanced datasets. However, this is a multifactorial problem and is associated with many challenges, such as historical discrimination, research design and accessibility [22]. We note that AI has the potential to address/mitigate bias without requiring such balanced datasets. A range of bias mitigation strategies have been proposed that either pre-process the dataset to make it less imbalanced, alter the training procedure or post-process the model outputs to reduce bias (27). We have recently proposed three algorithms to mitigate racial bias in CMR image segmentation: (1) train a CMR segmentation algorithm that ensures racial balance during training; (2) add an AI race classifier that helps the segmentation model to capture racial variations; and (3) train a different CMR segmentation model for each racial group. For more detail of these models, we refer to the reader to our previous work (14). All three proposed algorithms result in a fairer segmentation model that ensures that no racial group will be disadvantaged when segmentations of their CMR data are used to inform clinical management. Note that, compared to our previous work (14), in this paper we have excluded all subjects with cardiovascular disease to ensure that racial bias was not influenced by this factor.

### 4.4. Limitations

This study utilises the imaging cohort from the UK Biobank. UK Biobank is a long-term prospective epidemiology study of over 500,000 persons aged 40–70 years across England, Scotland, and Wales. Therefore, the data are geographically limited to the UK population, which might not reflect geographic, socioeconomic or healthcare differences among other populations. Manual analysis of CMR scans was performed by three independent centres using the same operating procedures for analysis. For the two centre, inter- and intra-observer variability between analysts was assessed by analysis of fifty, randomly-selected CMR examinations (17). However, one limitation of this study is that inter- and intra-observer variability was not assessed individually by race and sex. Also, this study is limited by the lack of diversity and relatively small sample sizes for certain racial groups and by the exclusion criteria for comorbid and pre-morbid conditions. The study only includes the following cardiovascular risk factors as confounders: hypertension, hypercholesteremia, diabetes and smoking. However, there are other clinically relevant risk factors such as sedentarism, alcohol consumption or stress that could potentially explain the bias found in our study. For instance, a previous study showed an association between RV size and living in a high traffic area (7). Finally, the outputs of this study are based on the ‘nnU-Net’ framework, and might not generalise to other DL-based segmentation models, although we note that nnU-Net is widely employed and was the best-performing model in a recent CMR segmentation challenge (6).

## 5. Conclusions

We have demonstrated that a DL-based cine CMR segmentation model derived from an imbalanced database has poor generalizability across racial groups and has the potential to lead to inequalities in early diagnosis, treatments and outcomes. Therefore, for best practice, we recommend reporting of performance among diverse groups such as those based on sex and race for all new AI tools to ensure responsible use of AI technology in cardiology.

## Data Availability

This research has been conducted using the UK Biobank Resource (application numbers 17806 and 2964). The UK Biobank data are available for approved projects from https://www.ukbiobank.ac.uk/

## Acknowledgements

This research has been conducted using the UK Biobank Resource (application numbers 17806 and 2964) on a GPU generously donated by NVIDIA Corporation. The UK Biobank data are available for approved projects from https://www.ukbiobank.ac.uk/.

## Abbreviations

AI: Artificial intelligence
CMR: Cardiac magnetic resonance
CVD: Cardiovascular diseases
DL: Deep learning
DSC: Dice similarity coefficient
ED: End diastole
EF: Ejection fraction
ES: End systole
LV: Left ventricle

## Clinical Perspectives

### Competency in medical knowledge

CMR can provide sensitive biomarkers for cardiac structure and function. However, analysis is time and labour intensive. DL can automate CMR analysis, but adequate sex and racial bias analysis is pivotal for clinical translation.

## Clinical competencies

### Competency in System-Based Practice

It is important to be aware that deep learning algorithms derived from imbalanced databases may be poorly generalizable and have the potential to reflect sex and racial inequalities.

## Translational Outlook

### Translational outlook 1

This is the first study to analyse the effect of sex and race on deep learning-based algorithms for CMR segmentation.

### Translational outlook 2

We show that current state of the art CMR segmentation pipelines can be racially biased and recommend adequate reporting of performance across racial/sex groups in the future.

---

**Central Illustration**

**Central Illustration: Fairness in automated CMR segmentation**

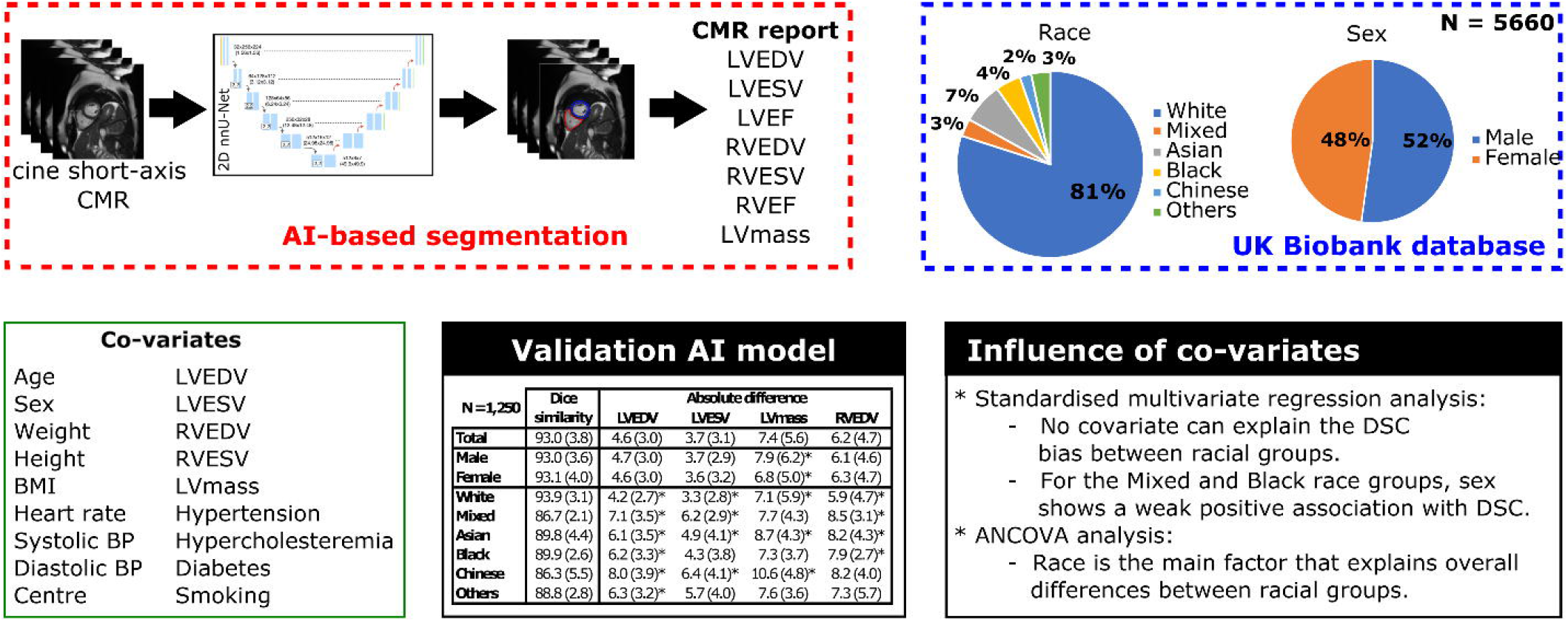

1 Office for National Statistics; National Records of Scotland; Northern Ireland Statistics and Research Agency (2017): 2011 Census aggregate data. UK Data Service (Edition: February 2017). DOI: http://dx.doi.org/10.5257/census/aggregate-2011-2

