## Supplemental appendix for "Fairness in Cardiac Magnetic Resonance Imaging: Assessing Sex and Racial Bias in Deep Learning-based Segmentation"

**Supplementary List 1. Exclusion criteria used for the selection of healthy volunteers from the UK Biobank database.**

**Age:** >75 years

**Medical conditions**: Adrenocortical insufficiency/Addison’s disease, Alcoholic liver disease/alcoholic cirrhosis, Anaemia, Angina, Ankylosing spondylitis, Anorexia/bulimia/other eating disorder, Antiphospholipid syndrome, Aortic aneurysm, Aortic regurgitation/incompetence, Aortic stenosis, Aplastic anaemia, Asthma, Atrial fibrillation, Atrial flutter, Bronchiectasis, Cardiomyopathy, Chronic obstructive airways disease, Clotting disorder/excessive bleeding, Connective tissue disorder, Crohn’s disease, Diabetes insipidus, Diabetic eye disease, Diabetic neuropathy/ulcers, Doctor diagnosed bronchiectasis, Emphysema, Emphysema/chronic bronchitis, Fibrosing alveolitis/unspecified alveolitis, Gestational diabetes, Gestational diabetes, Gestational hypertension/pre-eclampsia, Giant cell/temporal arteritis, Glomerulonephritis, Grave’s disease, Haemochromatosis, Haemophilia, Heart arrhythmia, Heart attack/myocardial infarction, Heart failure/pulmonary oedema, Heart valve problem/heart murmur, Heart/cardiac problem, Hereditary/genetic haematological disorder, Hyperaldosteronism/Conn’s syndrome, Hyperprolactinaemia, Hyperthyroidism/thyrotoxicosis, Hypertrophic cardiomyopathy, Hypopituitarism, Hypothyroidism / myxoedema, IgA nephropathy, Inflammatory bowel disease, Interstitial lung disease, Iron deficiency anaemia, Irregular heartbeat, Kidney nephropathy, Leg claudication/intermittent claudication, Liver failure/cirrhosis, Low platelets/platelet disorder, Lymphoedema, Microscopic polyarteritis, Miscarriage, Mitral regurgitation/incompetence, Mitral valve disease, Mitral valve prolapse, Monoclonal gammopathy/not myeloma, Myeloproliferative disorder, Myocarditis, Myositis/myopathy, Nephritis, Neutropenia/lymphopenia, Other respiratory problems, Pericardial effusion, Pericardial problem, Pericarditis, Peripheral vascular disease, Pernicious anaemia, Pleural effusion, Pleural plaques (not known asbestosis), Polycythaemia vera, Polymyalgia rheumatica, Polymyositis, Pulmonary embolism +/- DVT, Renal failure not requiring dialysis, Renal/kidney failure, Respiratory failure, Retinal artery/vein occlusion, Rheumatic fever, Sarcoidosis, Sick sinus syndrome, Sickle cell disease, Sjogren’s syndrome/sicca syndrome, Sleep apnoea, Stroke, Supraventricular tachycardia, Surgery/amputation of leg above the knee, Surgery/amputation of leg below the knee, Surgery/amputation of toe, Systemic lupus erythematosus, Transient ischaemic attack, Ulcerative colitis, Vasculitis, Wagner’s granulomatosis, Wolff-Parkinson-White syndrome.

**Medication:** Hormone replacement therapy.

**Symptoms:** Shortness of breath walking on level ground, Chest pain due to walking ceases when standing still, Chest pain when walking uphill or hurrying, Unable to walk up hills or to hurry.

**Supplementary Table 2.** **Associations between average DSC and each racial group**

|  | *Standardised beta-coefficients (95% CI)* | | | | | | |
| --- | --- | --- | --- | --- | --- | --- | --- |
|  | n | Model 3 - White | Model 3 - Mixed | Model 3 - Asian | Model 3 - Black | Model 3 - Chinese | Model 3 - Other |
| Age | 1024 | 0.03 (-0.02, 0.08) | -0.05 (-0.11, 0.00) | -0.05 (-0.11, 0.00) | -0.03 (-0.10, 0.03) | -0.04 (-0.11, 0.02) | -0.04 (-0.10, 0.02) |
| Sex | 1024 | 0.00 (-0.05, 0.06) | 0.09 (0.02, 0.16)* | 0.05 (-0.02, 0.12) | 0.10 (0.04, 0.18)* | 0.07 (-0.01, 0.14) | 0.08 (0.01, 0.15) |
| Weight | 1024 | 0.22 (-0.19, 0.66) | -0.01 (-0.58, 0.57) | 0.13 (-0.45, 0.75) | 0.27 (-0.27, 0.84) | -0.14 (-0.68, 0.43) | 0.01 (-0.60, 0.62) |
| Height | 1024 | -0.09 (-0.35, 0.17) | -0.01 (-0.36, 0.36) | -0.10 (-0.48, 0.29) | -0.18 (-0.52, 0.17) | 0.09 (-0.26, 0.45) | -0.04 (-0.42, 0.36) |
| BMI | 1024 | -0.12 (-0.47, 0.20) | 0.06 (-0.40, 0.54) | -0.07 (-0.57, 0.41) | -0.19 (-0.65, 0.25) | 0.17 (-0.28, 0.61) | 0.03 (-0.46, 0.53) |
| HR | 1024 | 0.03 (-0.01, 0.07) | 0.03 (-0.02, 0.09) | 0.03 (-0.02, 0.09) | 0.01 (-0.04, 0.06) | 0.03 (-0.02, 0.09) | 0.03 (-0.03, 0.08) |
| SBP | 1024 | -0.02 (-0.07, 0.04) | -0.01 (-0.07, 0.05) | -0.02 (-0.09, 0.04) | -0.02 (-0.08, 0.04) | -0.03 (-0.09, 0.03) | -0.03 (-0.09, 0.03) |
| DBP | 1024 | -0.03 (-0.08, 0.01) | -0.01 (-0.07, 0.05) | -0.01 (-0.08, 0.04) | -0.01 (-0.07, 0.05) | -0.03 (-0.08, 0.03) | -0.01 (-0.06, 0.05) |
| LVEDV | 1024 | -0.03 (-0.25, 0.17) | -0.15 (-0.43, 0.14) | -0.16 (-0.45, 0.11) | -0.15 (-0.42, 0.14) | -0.13 (-0.41, 0.13) | -0.14 (-0.41, 0.13) |
| LVESV | 1024 | -0.06 (-0.20, 0.07) | -0.07 (-0.27, 0.11) | -0.06 (-0.25, 0.14) | -0.07 (-0.26, 0.11) | -0.08 (-0.26, 0.12) | -0.07 (-0.25, 0.12) |
| RVEDV | 1024 | 0.14 (-0.06, 0.36) | 0.25 (-0.07, 0.55) | 0.26 (-0.04, 0.58) | 0.26 (-0.06, 0.58) | 0.22 (-0.10, 0.54) | 0.23 (-0.08, 0.52) |
| RVESV | 1024 | -0.11 (-0.27, 0.03) | -0.16 (-0.38, 0.05) | -0.17 (-0.39, 0.04) | -0.18 (-0.40, 0.03) | -0.16 (-0.38, 0.06) | -0.15 (-0.35, 0.07) |
| LVmass | 1024 | -0.02 (-0.09, 0.04) | -0.01 (-0.08, 0.07) | 0.02 (-0.07, 0.10) | -0.06 (-0.14, 0.02) | 0.00 (-0.09, 0.08) | -0.01 (-0.09, 0.08) |
| Diabetes | 1024 | 0.08 (-0.10, 0.25) | 0.11 (-0.10, 0.31) | 0.12 (-0.10, 0.34) | 0.19 (-0.01, 0.41) | 0.15 (-0.07, 0.36) | 0.14 (-0.08, 0.36) |
| Hypertension | 1024 | 0.06 (0.01, 0.11) | 0.03 (-0.02, 0.09) | 0.04 (-0.02, 0.10) | 0.04 (-0.01, 0.10) | 0.03 (-0.03, 0.09) | 0.05 (-0.01, 0.11) |
| Hypercholesterolemia | 1024 | -0.01 (-0.05, 0.04) | 0.02 (-0.04, 0.08) | 0.00 (-0.06, 0.06) | 0.00 (-0.06, 0.06) | 0.01 (-0.05, 0.07) | 0.01 (-0.05, 0.07) |
| Smoking | 1024 | 0.00 (-0.04, 0.04) | -0.01 (-0.06, 0.04) | 0.00 (-0.05, 0.05) | -0.01 (-0.05, 0.05) | 0.00 (-0.06, 0.04) | -0.01 (-0.07, 0.04) |
| Centre | 1024 | 0.14 (0.08, 0.20) | -0.25 (-0.32, -0.18) | -0.26 (-0.34, -0.18) | -0.24 (-0.31, -0.17) | -0.26 (-0.33, -0.20) | -0.26 (-0.33, -0.19) |
| Racial group | 1024 | -0.74 (-0.80, -0.69)** | 0.23 (0.19, 0.27)** | 0.17 (0.11, 0.21)** | 0.24 (0.20, 0.28)** | 0.21 (0.17, 0.24)** | 0.18 (0.15, 0.21)** |

Standardized regression beta-coefficients and CI, representing the z-score change in variables with increasing DSC. Each model is for a comparison between each racial group vs all other racial groups (i.e. Model 3 is the regression analysis for the White group vs the combination of Mixed, Asian, Black, Chinese and Other groups). LV: left ventricle, EDV: end-diastolic volume, ESV: end-systolic volume, SBP: systolic blood pressure, DBP: diastolic blood pressure; CI: confidence interval. Model 3 is the multivariate linear regression per racial group adjusted for sex, height, weight, blood pressure at scan-time, heart rate at scan-time, LVEDV, LVESV, RVEDV, RVESV, LVmass, diabetes, hypertension, hypercholesterolemia, smoking and centre. * *p<.01, ** p<.001, *** p<.0001.*

**Supplementary Table 3. Results of the one-way ANOVA and ANCOVA.**

| (a) Model 4 (1-way ANOVA) | | | |
| --- | --- | --- | --- |
|  | F | Sig. | η^2^ |
| Racial group | 219.43 | 0.00 | 0.47 |
| (b) Model 5 (ANCOVA) | | | |
|  | F | Sig. | η^2^ |
| Age | 1.51 | 0.13 | 0.00 |
| Sex | 0.61 | 0.22 | 0.00 |
| Weight | 0.18 | 0.43 | 0.00 |
| Height | 0.00 | 0.67 | 0.00 |
| BMI | 0.01 | 0.98 | 0.00 |
| HR | 1.88 | 0.93 | 0.00 |
| SBP | 0.31 | 0.17 | 0.00 |
| DBP | 2.32 | 0.58 | 0.00 |
| LVEDV | 0.03 | 0.13 | 0.00 |
| LVESV | 1.05 | 0.87 | 0.00 |
| RVEDV | 0.95 | 0.30 | 0.00 |
| RVESV | 1.74 | 0.33 | 0.00 |
| LVmass | 1.87 | 0.19 | 0.00 |
| Diabetes | 0.97 | 0.17 | 0.00 |
| Hypercholesterolemia | 0.03 | 0.33 | 0.00 |
| Hypertension | 4.85 | 0.85 | 0.00 |
| Smoking | 0.04 | 0.03 | 0.00 |
| Centre | 32.18 | 0.84 | 0.03 |
| Racial group | 167.48 | 0.00 | 0.41 |

Outcomes of the 1-way ANOVA (model 4) and the ANCOVA (model 5) adjusted for sex, height, weight, blood pressure at scan-time, heart rate at scan-time, LVEDV, LVESV, RVEDV, RVESV, LVmass, diabetes, hypertension, hypercholesterolemia, smoking and centre. LV: left ventricle, EDV: end-diastolic volume, ESV: end-systolic volume, SBP: systolic blood pressure, DBP: diastolic blood pressure.
